# Feasibility of Establishing a Return-To-Work Protocol Based on COVID-19 Antibodies Testing

**DOI:** 10.1101/2020.09.03.20187823

**Authors:** Jennifer Okungbowa-Ikponmwosa, Yijia Mu, Gerard Job

**Affiliations:** Emergency department, Jackson Memorial Hospital, Miami Florida; Miami Dade Fire, Air and Ocean Rescue, Miami Florida

**Keywords:** COVID-19, SARS-CoV-2, Rapid IgM-IgG Combined test, Point-of-Care Testing, Return-to-Work protocol

## Abstract

**Introduction:** Prior to the diagnosis of the first SARS-CoV2 patient in Florida, the Miami Dade Fire Rescue developed and implemented its return-to-work protocol based on guidelines from the CDC and Florida Fire Chiefs Association. As of February 17, 2020, all asymptomatic employees exposed to PCR-confirmed positive SARS-CoV2 individuals would be excluded from work for 14 days and report absence of symptoms to a delegated supervisor every 24 hours. We postulated that if COVID-19 transmission rate continues at the current rate in the absence of systemic vaccination strategy for SARS-CoV2, then a safer and more efficient return-to-work policy is needed for exposed first responders who are identified as low-risk for disease transmission.

**Objectives:** We sought to establish a safe and shortened return-to-work protocol to maintain our workforce. We evaluated the utility of serological antibody testing in predicting negative seroconversion of first responders at 7 days post low-risk exposure to confirmed COVID-19 individuals.

**Methods:** All exposed, asymptomatic employees underwent serology testing for SARS-CoV2 one week after the initial exposure. Participants who were serologically negative had follow-up RT-PCR within 24 hours and serology testing 14 days after the initial serological test.

**Results:** Overall, of the 71 firefighters who have had documented exposures to SARS-CoV2 positive individuals in the fire rescue agency, 41 of 71 had initially negative serology studies. Of the 41 patients with negative serology studies, 20 voluntarily underwent confirmatory PCR testing within one day after serology testing and all 20 participants were negative. Subsequently, out of the 20 participants who underwent serology and PCR testing, 10 participants followed up and underwent repeat serology testing 14 days after exposure and all 10 participants had negative repeat serology tests. The other ten who chose not to retest remained asymptomatic 14 days after exposure.

**Conclusions:** Although serology testing has limitations, it correlated with negative prediction of disease in low-risk participants with exposures in this study. Serology testing may offer a feasible, alternative return-to-work strategy for fire agencies.

## Introduction

The SARS-CoV2 (COVID-19) outbreak has been declared a pandemic by the World Health Organization on March 11, 2020 and is the first pandemic since the H1N1 influenza outbreak. The virus was first detected in December 2019 in Wuhan, China^1^ and quickly spread to other continents within 2–3 months. Similar to other beta-coronaviruses infection, SARS-CoV2 triggers humoral and adaptive cellular immunity in humans leading to its antibody-mediated neutralization and clearance. Prior studies of SARS-CoV1 have demonstrated that specific antibodies can be detected as early as 0 to 7 days after symptoms onset with conferred immunity up to 2 years after infection^2^. More recent, early serological COVID-19 studies had led to the development of rapid lateral flow assay tests which detect antibodies to SARS-CoV2 proteins and provide an indirect method of detecting the COVID-19 infection. It is uncertain exactly when antibodies can first be detected in the bloodstream but the American Society for Microbiological at its international Summit in March, 2020 indicated that the majority of patients seroconvert between 7–11 days post-exposure.^3^ While RT-PCR remains the gold standard for diagnosing SARS-CoV2 infection currently, the test is both time- and resource-intensive, requires specialized supplies, expensive instrumentation and laboratory-trained technicians to perform the test. Serological antibodies testing offers quicker, more logistically feasible alternatives for designing a rapid testing ‘return-to-work protocol’.

With the increased incidence of exposure to COVID-19, the Miami Dade Fire Rescue, a large county-based fire agency, sought to implement a safe and efficient return-to-work policy for its first responders who were considered to have had a low-risk exposure and were asymptomatic. In our fire agency, a low-risk exposure was defined as any worker who had less than 15 minutes interactions with patients with COVID-19 or who had prolonged exposure to patients with COVID-19 but were wearing PPE including a respirator and eye protection at the time of exposure. The return to work policy was created and implemented on February 17, 2020. It mandated that all asymptomatic individuals exposed to PCR-confirmed positive COVID-19 patients would be excluded from work for 14 days and self-monitored with a delegated supervisor. Our policy largely mirrored the CDC guidelines and the Florida Fire Chiefs Association guidelines. Since the implementation of the policy, there have been significant financial and logistical consequences with increased burden on non-exposed first responders. Due to the consequences of the protocol, it was imperative to develop a new policy to safely reduce the 14-day work exclusion for exposed asymptomatic firefighters while maintaining the safety of other personnel. Thus, we considered the role of serological testing in conjunction with PCR to assess the feasibility of revising the return to work protocol.

## Methods

As part of a Public Health strategy, Miami Dade County in collaboration with the University of Miami initiated a county-wide antibodies testing to determine the prevalence of COVID-19 in their community (SPARK-C). This study included 2038 sworn firefighters (79% males and 53% white) from Miami Dade Fire Rescue ranging in age from 20 −70 years. They were voluntarily tested with BioMedomics COVID-19 IgM-IgG rapid Test which used recombinant antigen (MK2010227), a receptor binding domain SARS-CoV2 Spike Protein (RBD-S) to detect antbodies.^4^

Inclusion criteria were firefighters in Miami Dade County who were exposed to an individual who had positive antibody or PCR testing for COVID-19 and were asymptomatic.

Between the period of April 17 to May 1, 2020; all asymptomatic individuals that were exposed to a serological positive individual or a laboratory confirmed RT-PCR positive individual were offered testing with the BioMedomics COVID-19 IgM-IgG test 7 days after the day of most recent exposure and RT-PCR within 24 hours to confirm the diagnosis..

Participants who were serologically negative for antibodies were asked to have a follow up RT-PCR within 24 hours and a repeat rapid antibodies test at 14 days. Data gathered was then used to calculate the negative predictive value in asymptomatic patients who had negative serology and PCR. While the positive predictive value was also calculated for patients who were asymptomatic with positive serology and positive PCR.

All positive individuals were excluded from work.

## Results

From the SPARK-C study and infection control office, 71 individuals were identified that were exposed to serological antibody positive co-workers or RT-PCR positive patients and were all tested with rapid COVID-19 test. Approximately 60% tested negative irrelevant of their gender (table1). Twenty (20) of the forty-one (41) negative rapid COVID-19 individuals volunteered to be tested with RT-PCR and were all negative. This indicates 100% negative predictable value (see table1 and figure 1). Ten (10) followed up for the repeat rapid antibodies 14 days after the initial test and again tested negative. The other ten chose not to retest but remained asymptomatic 14 days after exposure. Twenty-six (26) of the thirty (30) positive rapid COVID-19 individuals were negative when tested with RT-PCR indicating a dismal positive predictable value of 13% (see Figure 1 and Table 1).

**Figure 1:**
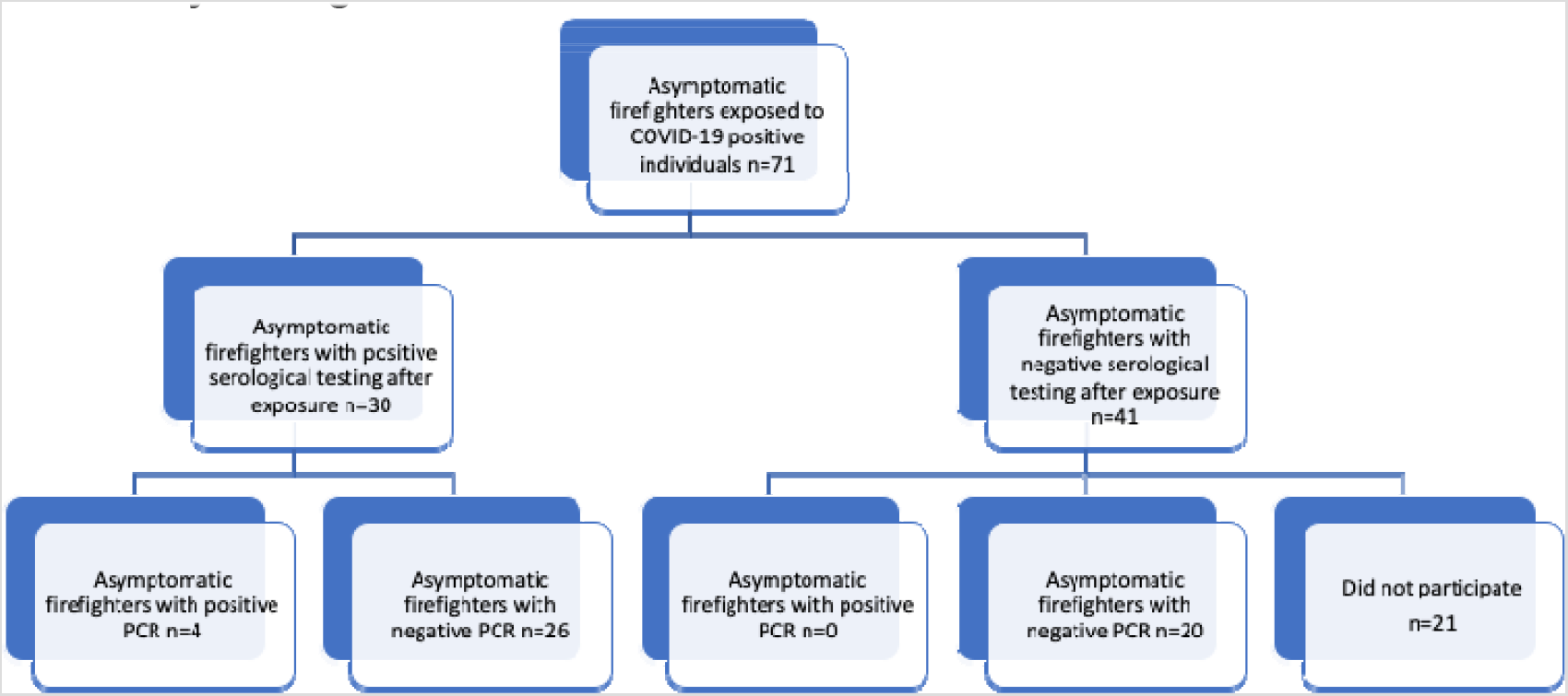
Results of asymptomatic firefighters who underwent serology testing and confirmatory testing with PCR

**TABLE 1:** SEX AND AGE DATA

| SEX | TOTAL n=71 | SEROLOGY<br>NEGATIVE | NEGATIVE<br>PREDICTABLE<br>VALUE (TRUE<br>NEGATIVE) | POSITIVE<br>PREDICTABLE<br>VALUE (TRUE<br>POSITIVE) |
| --- | --- | --- | --- | --- |
| MALE | 61 (86%) | 35 (57%) | 16/16 (100%) | 4/26 (19%) |
| FEMALE | 10 (14%) | 6 (60%) | 4/4 (100%) | 0/4 (0%) |
| AGE (YEARS) |  |  |  |  |
| 20-29 | 8 | 3 (38%) | 2/2 (100%) | 1/5 (20%) |
| 30-39 | 21 | 16 (76%) | 7/7 (100%) | 0/5 (0%) |
| 40-49 | 26 | 14 (54%) | 6/6 (100%) | 1/12 (8%) |
| 50-59 | 13 | 7 (54%) | 4/4 (100%) | 2/6 (33%) |
| 60-69 | 3 | 1 (33%) | 1/1 (100%) | 0/2 (0%) |

## Discussion

Covid-19 in low risk patients (asymptomatic exposed) can be safely excluded with serological antibodies testing after 7 days and maybe used to establish ‘return to work’ protocols. In our study, low-risk patients can be defined per CDC guidelines to include any healthcare personnel who had brief interactions with patients positive for COVID-19 or those who had prolonged exposure to patients positive COVID-19 but were wearing PPE including face masks or a respirators at the time of exposure. While RT-PCR remains the gold standard for diagnosing SARS-CoV2 infections currently, it is time-intensive and resource-scarce, requiring specialized supplies, expensive instruments and laboratory trained technicians.

The use of serological antibodies as surrogates for SARS-CoV2 infections show promise for indirectly identifying COVID infections. Recent early studies have indicated that seroconversion may occur 7–11 days post exposure^3^ and IgM has been found to be positive as early as 4 days after symptoms onset^5,6^. Since the average incubation period is 4–5 days^7^, it seems reasonable to assume that asymptomatic individuals may be classified low risk for infection and return to work, if they remain asymptomatic a week after initial exposure. This was what we set out to confirm when we tested our exposed crew members with rapid serological antibodies tests 7 days after being exposed to a laboratory confirmed COVID-19 positive person or a serological IgM+/IgM-IgG+ co-worker. This time period of self-monitoring was used to eliminate false negatives that might be possible before seroconversion.

From our study, we were able to deduce that the negative predictive value in asymptomatic firefighters who had low-risk exposure to COVID-19 with negative serology and PCR was 100% at 7 days and 21 days after initial exposure. Therefore, there may be utility in using rapid serology tests as an initial screening tool to rule out COVID-19 in low risk patients. Also, we can further consider using this test as an adjunct for developing a protocol that would allow employees to return to work quicker. Specifically, our results showed that it may be feasible to safely develop a return to work protocol whereby workers could be able to return to work 7 days earlier than the current recommendations set out by the CDC for workers who have had low-risk exposure to COVID-19.

Initial limitations to our study include small sample sizes. Although the inclusion criteria allowed for the eligibility of 71 participants out of a group of 2038 firefighters, 21 from the negative serology group did not enrol in the PCR testing following serological testing. It is unclear whether or not a lower number of participants in this group lead to a high negative predictive value. Nevertheless, given negative initial serology it is unlikely that this was the case. Further studies could investigate how return to work protocols can be developed for employees who have serologic and PCR testing with discordant results.

Other limitations involve the utility of serological markers. Many questions remain unanswered including how soon does seroconversion occur after positive exposure, how accurate is antibodies testing for identification of acute SARS-CoV2 infection vs convalescence from prior infection, the implication of positive serology markers for the SARS-COV-2 reinfection and duration of immunity. Most pressing of these concerns is serology test validation. To date, there are more than a 100 commercially available serology tests for SARS-CoV2 in the United States with variable reliability and accuracy for the diagnosis of SARS-CoV2. While the Food and Drug Administration (FDA) has begun overseeing a validation process for serological tests, it is still unclear when this investigation will be completed. Second it is still unclear whether specific antibody level as measured by serology studies indicates any clinically relevant immunity from future SARS-CoV2 infections. These questions along with the increasing number of unvalidated serology tests without Emergency Use Authorization have resulted in the CDC and WHO recommendation for research only purposes for these tests currently.

Although these limitations exist, implementing a serology-based return-to-work policy may be practical and economically reasonable for a large county fire rescue agency. In our study, it reduced our period of exclusion from work by 50% and ensured that staff would be available in contingency and crisis surge situations. Therefore, the utility of serology-based return-to-work policy in anticipation of further validation studies from the FDA and other health organizations may be evaluated in a future prospective study as commercial serology kits become readily available.

## Conclusions

This study aimed at creating a feasible return-to-work policy that prioritizes worker’s safety while reducing the financial burden incurred from personnel availability, logistical limitations, and decreased productivity imposed on EMS systems during the SARS-CoV2 pandemic. Although a number of limitations exist to using serology tests currently; in instances where PCR-based studies are not readily available, serology studies may be considered as a viable alternative strategy to quickly and effectively risk-stratify SARS-Cov2 low-risk exposures in first responders. In Miami-Dade County, the use of serology tests identified and predicted safe return-to-work status for rescue personnel who had low-risk exposure to SARS-CoV2 with negative follow-up serology and PCR confirmations. Further prospective validation studies are needed to evaluate the use of serology-based testing in low-risk EMS cohorts. Until PCR-based studies have become readily available and quickly accessible for first responders, serology-based studies offer a feasible and reasonable alternative strategy for a return-to-work protocol when combined with clinical suspicions in the SARS-CoV2 pandemic.

## Data Availability

Accompanying data will be made available upon request.

